# Functional Rating Scales in Spinal and Bulbar Muscular Atrophy: A Systematic Review, Meta-Analysis and Critical Appraisal of their Measurement Properties

**DOI:** 10.1101/2021.05.30.21258087

**Authors:** Agessandro Abrahao, Liane Phung, Eliza Freitas, Cornelia M. Borkhoff, Lorne Zinman

## Abstract

Tracking disease progression and treatment effect of spinal bulbar muscular atrophy, or Kennedy’s disease, is challenging given its slowly progressive nature. To achieve success in SBMA clinical trials, a reliable, responsive, and validated patient-reported motor function scale must capture progression of SBMA-specific motor dysfunction. Here, we conducted a systematic review, meta-analysis, and appraisal of core measurement properties of the SBMA functional rating scale (SBMAFRS). We established that the SBMAFRS has satisfactory internal consistency, inter-rater reliability, and construct validity for measuring progressive motor dysfunction over similar neurodegenerative motor function scales but inadequate sensitivity to change over time. Further development to validate and improve the SBMAFRS’ ability to capture longitudinal responsiveness in larger cohorts is warranted.

## 1. INTRODUCTION

Spinal and bulbar muscular atrophy (SBMA) or Kennedy’s disease is an X-linked, adult-onset multisystem disorder affecting the lower motor neurons, sensory nerves, and muscle fibres, along with features of partial androgen insensitivity. The disease is caused by a CAG trinucleotide repeat expansion in the androgen receptor gene [1] and has a prevalence of 1 in 40,000 men [2]. To date, there is no effective treatment to halt or cure the disease.

Tracking disease progression or quantifying a small treatment effect in longitudinal SBMA studies is challenging given the slowly progressive nature of the disease. In contrast to amyotrophic lateral sclerosis (ALS) – the fastest motor neuron disease, the motor examination may remain relatively stable over one to two years in SBMA, which is a common duration chosen for SBMA pivotal trials. In randomized, placebo-controlled SMBA trials that reported negative outcomes [3–5], a small effect size may have been missed by insensitive endpoint measures of motor function.

To achieve success in clinical trials of rare diseases such as SMBA, a reliable primary endpoint of disease progression that is responsive to change over a relatively short interval is warranted. While surrogate biomarkers have not been fully validated in SBMA, existing patient-reported motor function scales, such as the revised ALS functional rating scale (ALSFRS-R) and quantitative myasthenia gravis (QMG), have been recycled as outcome measures in SBMA studies [5–8]. However, an optimal patient-reported functional outcome scale for an evaluative purpose [9] should be SBMA-specific and thoroughly quantify the core concept of progressive motor dysfunction in multiple domains (i.e., bulbar, cervical, thoracic and lumbosacral regions). The items of the scale should have enough gradation to capture small changes in function over just 6 to 12 months to be utilized in smaller, shorter and less costly clinical trials. For SMBA, the assessment of extra-motor functional loss is not essential in a measurement instrument of longitudinal change as these features tend to remain stable over time.

In this study, we conducted a systematic review and meta-analysis of SBMA-specific, patient-reported functional scales and appraised whether their sensibility and measurement properties fit the conceptual framework of small-magnitude longitudinal motor dysfunction to serve as outcome measures in short-term clinical trials or cohort studies.

## 2. METHODS

### 2.1 Literature Search

A literature search was conducted on three public databases: PubMed, Medline, and Embase (1966-September 28, 2020), to identify potential instruments that would fit the proposed measurement concept. Search strategy followed the Consensus-based Standards for the Selection of Health Measurement Instruments (COSMIN) recommendations [10] and included the following key elements: 1) construct of interest: functional OR function OR weakness OR loss of function OR symptom OR disease progression OR functional loss; 2) target population: Kennedy’s disease[MeSH term] OR “Kennedy’s disease” OR “spinal and bulbar muscular atrophy” OR “spinal bulbar muscular atrophy”; 3) type of measurement instrument: scale OR index OR questionnaire OR instrument; 4) measurement properties: COSMIN inclusion and exclusion filters [10]. Detailed search strategy is described in the **Supplementary Table 1**.

### 2.2 Data Extraction

Two independent extractors (AA and EF) initially screened the article titles and abstract for relevant entries. Full-text from candidate articles were reviewed and studies reporting the development, measurement properties and validation of motor function scales in SBMA were included. Extracted data included scale denomination, scale development (item generation, selection and reduction, and scoring) and the following measurement properties: sensibility, content validity, face validity, feasibility, reliability, construct validity, and responsiveness.

### 2.3 Statistics

Meta-analyses of single-group means for age at enrollment, age at SBMA onset, disease duration, CAG repeat length, and SBMAFRS scores were estimated using random-effects models with inverse variance method and DerSimonian-Laird τ^2^ estimator. Pooled mean value across studies, standard deviation (SD), and 95% confidence interval (95%CI) of the mean were reported for each variable. Meta-regression of mean SMBAFRS values was modelled with mean age, mean age at disease onset, disease duration and mean CAG nucleotide repeats as moderators with restricted maximum likelihood τ ^2^estimator. Analyses were computed using R 3.6.2 (meta and metafor packages).

## 3. RESULTS

### 3.1 Identified instruments

Eighteen articles were identified and four were included (**Supplementary Figure 1**). Two SBMA-specific, patient-reported motor functional scales were identified: the SBMA Functional Rating Scale (SBMAFRS) [11–13] and the Kennedy’s disease 1234 scale (KD 1234 scale) [14]. For the purpose of this review, only the SBMAFRS was appraised and included in the meta-analysis as three studies had tested the scale in multiple cohorts, and in multiple languages (**Table 1**).

**Table 1.** Evidence of reliability and validity of included articles

| **SBMAFRS** | **Study design** | **Sample size** | **Measurement property** | **Findings** |
| --- | --- | --- | --- | --- |
| Hashizume et al. 2015 | Prospective Cohort | 41 cases/41 controls | **Reliability**<br>- test–retest reliability: ICCs with a one-way mixed effect model;<br>- inter-rater reliability: ICCs with a two-way mixed effect model | Excellent Intra-rater agreement (95% CI): 0.910 (0.811–0.958).<br>Inter-rater reliability was high (95% CI): 0.797 (0.684–0.870), although detailed analysis of the ICCs for each item indicated lower values than for intra-rater reliability. |
|  |  |  | **Validity**<br>- internal consistency: Cronbach’s alpha;<br>- inter-scale correlations<br>- convergent and discriminate construct validity | Internal consistency was satisfactory, with Cronbach’s alpha values ranging from 0.700 to 0.822.<br>Inter-rater reliability and internal consistency were similar to those of the SBMAFRS-J, although the ICC for the breathing- related subscale of the SBMAFRS-E was better than that of the SBMAFRS-J.<br>The bulbar-related subscale of the SBMAFRS correlated best with that of the ALSFRS-R and Norris Bulbar Score, and worst with the lower limb- or respiration-related subscale of the ALSFRS-R and Limb Norris Score. Conversely, the lower limb-related subscale of the SBMAFRS correlated best with that of the ALSFRS-R and Limb Norris Score, and worst with the bulbar-related subscale of the ALSFRS-R and Norris Bulbar Score. |
| Querin et al. 2016 | Prospective cohort | 60 patients | **Reliability**<br>- test–retest reliability, Spearman’s rank order correlation coefficient (Spearman’s rho)<br>- the percentage of perfect agreement was calculated to measure the level of agreement on item level. | Spearman’s rank order correlation coefficients showed strong positive correlations which were statistically significant for subscores, both for single items and in total.<br>Perfect agreement between test and retest on subscale level showed high test–retest reliability: 76% (total score) |
|  |  |  | **Validity**<br>- Convergent validation was calculated by comparing the scores achieved in items of the SBMAFRS with the corresponding ones of the ALSFRS | Cronbach’s alpha coefficient was adequate (total raw alpha = 0.85, standardized alpha = 0.84). |
| Dahlqvist et al. 2018 | Prospective cohort | 29 patients | Disease progression investigation using MRI to evaluate changes in muscle fat content and stationary dynamometry to assess changes in muscle strength. Disease progression was also investigated with the SBMAFRS, bulbar rating scale, 6-minute walk test and blood samples among others. | The SBMAFRS and BRS scores did not change: SBMAFRS score was 76% of normal at baseline and 75% of normal at follow-up, and the BRS score was 91% of normal at baseline and 89% of normal at follow-up. |

SBMAFRS: The Spinal and Bulbar Muscular Atrophy Functional Rating Scale; KD 1234: Kennedy's disease 1234 scale; ICC: intra-class correlation coefficients; SBMAFRS-J: Japanese version; SBMAFRS-E: English version

### 3.2 Sensibility of the SBMAFRS

The appraisal of the sensibility and clinical usefulness of the SBMAFRS followed the framework proposed by Bombardier & Tugwell, 1987[15] and modified by Buchbinder et al. 1996 [16].

#### 3.2.1 Purpose, Population and Setting

The SMBAFRS was developed as an evaluative scale probing the concept of multi-domain motor dysfunction for clinical trials and cohort studies. This scale does not measure extra-motor features of SBMA.

#### 3.2.2 Content and Face Validity

The SBMAFRS measures motor disability for activities of daily living (ADLs) and physical examination of motor function. It has an adequate readability and understandability, without double negative, double-barreled or false premise questions.

The 14-item questionnaire was designed with the assumption of five functional domains: bulbar subscale (five items), upper limb subscale (two items), truncal subscale (four items), lower limb subscale (two items), and breathing subscale (one item). Each item has five response options scoring 0 (worst) to 4 (normal). The sum score ranges 0 to 56. The full instrument can be extracted from Hashizume et al. 2015 [11].

The non-uniform item distribution with relatively more items measuring bulbar and trunk-related tasks compared to breathing and appendicular function enhances the potential responsiveness to longitudinal change and face validity of the SBMAFRS. Compared to the most utilized ALS functional scale (ALSFRS-R) [17] which has equal item distribution among bulbar, limb and breathing domains, the SBMAFRS is weighted on phenotypic features that are more likely to change over time. Weakness in SBMA is slow in progression over years and affects predominantly proximal muscles, while weakness in typical ALS cases spreads throughout the body within months. Severe respiratory failure – a frequent cause of death in ALS – is uncommon in SBMA and, therefore, is measured by only one item in the SBMAFRS questionnaire.

#### 3.2.3 Feasibility

The original SBMAFRS was developed in Japanese and translated into English (SBMAFRS-E) and Italian (SMAFRS-I) [13], following standard cross-cultural validation methods [18]. The successful implementation of the SBMAFRS instrument in two Japanese[11], one American[11] and two Italian[13] centres during the validation stage, along with a published rating algorithm, are encouraging for the feasible use of the scale as an endpoint in multicentre trials and cohort studies.

Remote administration of the SBMAFRS is a limitation and has not been validated. The instrument incorporates parameters obtained from physical examination, such as puffing cheeks (evaluating facial weakness), push-ups, tongue atrophy and motor tasks. Phone administration of the SBMAFRS is impracticable; however, future studies can compare SBMAFRS scores obtained during in-person visits to videoconferencing administration. Future validation of video-captured SBMAFRS scores may reduce missing data from participants who would drop out from in-person study visits due to immobility. Indeed, prior SBMA drug trials showed a dropout rate of 10 to 20% [4, 8, 19].

### 3.2 Development stage of the SBMAFRS

#### 3.3.1 Item generation

The development of the SBMAFRS included items adapted from scales tailored for patients with ALS, such as the ALSFRS-R [17] and the modified Norris Scale [20]. The 12-item ALSFRS-R measures the motor disability and impairment of ADLs related to bulbar weakness, fine- and gross-motor dysfunction, and respiratory failure. The Norris Scale, published in 1974 [20], is one of the oldest ALS measurement instruments. The modified version incorporate the Limb Norris Score with 21 items and the Norris Bulbar Score with 13 items [21].

A limitation of using ALS-related items in a slowly progressive disease such as SBMA is the potential compromise in short-term responsiveness. Given that ALS is rapidly progressive, changes in ALSFRS-R scores can be detected within 1 to 3 months for most patients. Indeed, prior SBMA cohort and interventional studies that have applied the ALSFRS-R [5–8] and the modified Norris Scale [6, 22, 23] as endpoints reported limited responsiveness in the short-term. Detectable between-group changes in these scales required much longer follow up in these studies (up to 84 months [22]).

Original items were also generated for the SBMAFRS to better reflect the slower SBMA motor phenotype as compared to ALS. Items measuring the ability to puff up the cheeks, rise from a sitting or supine position, ability to bow, the presence of drooling and tongue atrophy were proposed.

#### 3.3.2 Item Selection

From the candidate item pool, a panel of seven SBMA experts selected the 14 most relevant entries, modified the five response categories per item, and eliminated redundancy. Selection criteria followed a blend of explicit judgement and statistical approach, whereby published score distribution of ALSFRS-R and Norris Scale items [4, 6, 8, 19] were considered to identify relevant items.

Reporting of the item selection process was unclear regarding the method of expert panel interaction and approach to achieve consensus. Also, the lack of a patient focus group and stakeholder input on relevant item selection is an important limitation as SBMA patients may attribute discordant weights to their perceived deficits compared to physicians’ opinions.

### 4. Cross-sectional and Cohort Validity Studies of SBMAFRS

Cross-sectional and cohort studies have applied the Japanese and translated SBMAFRS questionnaires in 121 Japanese [11], 15 American [11], and 60 Italian [13] patients. The SBMAFRS global score was normally distributed, without a reported floor or ceiling effect in these studies. Male healthy controls scored near the ceiling in the SBMAFRS (55.9±0.4 points, n=41) [11].

Cronbach’s alpha internal consistency and confirmatory factor analysis were tested in the Japanese, American and Italian patient populations (**Table 1**). Cronbach’s alpha values were satisfactory (0.7 to 0.85) in the Japanese and Italian cohorts [11, 13], but suboptimal values (below 0.7) were extracted in the American population [11], which may be related to the small sample size. The assumption of five independent SBMAFRS subscales was not validated by the confirmatory factor analysis using oblique and varimax rotation in the Japanese and Italian cohorts. These statistical tests extracted three domains instead: 1. trunk and lower limb domain; 2. bulbar and upper limb domain; and 3. breathing domain [11, 13]. This discrepancy between the assumed versus data-driven domains may be related to the myotomal nature of SBMA disease progression.

The SBMAFRS subscales had significant Spearman’s correlation with the ALSFRS-R subscales (coefficient ≥0.7), except for a modest correlation between the breathing subscales (coefficient 0.5 to 0.7). SBMAFRS limb subscales also had good correlation with 6-minute walking test and manual muscle strength test [11, 13]. These data establish the appropriateness of the SBMAFRS convergent and divergent construct validity.

[14][14]

### 5. Meta-analysis

In a meta-analysis of SBMAFRS validation studies (**Figure 1**), the pooled mean age at enrolment was 57 years (95%CI 54, 59) and mean age at onset was 43 years (95%CI 42, 45). Mean CAG repeat length was 47 (95%CI 45, 49) and mean disease duration was 14 years (95%CI 11, 17). These key demographic and genotypic characteristics were heterogenous with high I^2^ across studies, except for mean age at disease onset.

**Figure 1.**
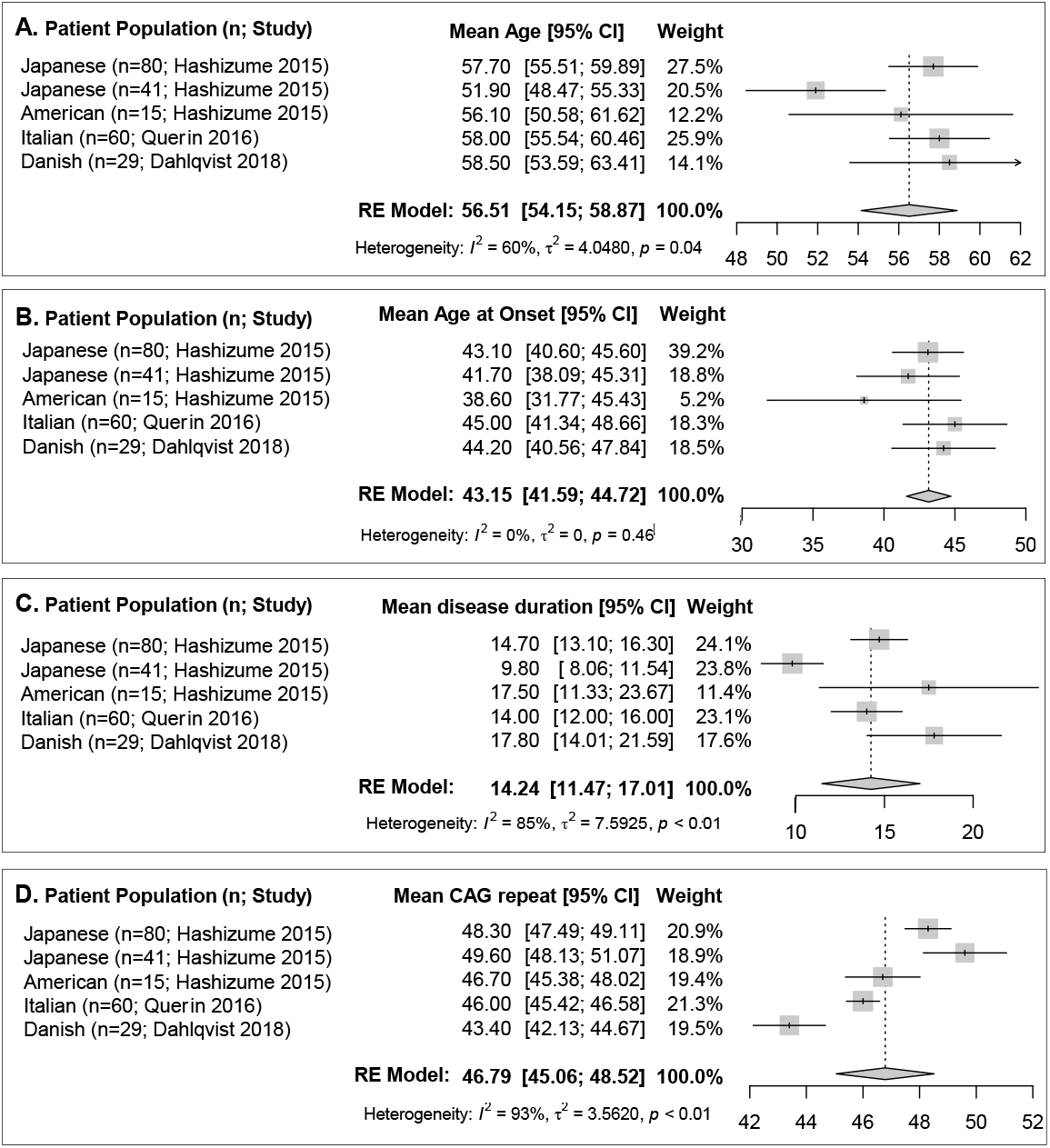
Forest plots of pooled mean age (A), mean age at onset (B), mean disease duration (C), and mean CAG repeat lengths (D) of five populations from three published studies.

The pooled mean SBMAFRS global score was 40.4 (95%CI 37, 44) with significant between-study heterogeneity (**Figure 2**). In meta-regression models, almost 25% of the variability in SBMAFRS scores across studies was attributed to age at disease onset, where younger onset age associated with lower scores (R^2^ 24.6%, p <0.001, **Figure 3B**). Mean SBMAFRS scores had a non-significant decline as disease duration or the number of CAG trinucleotide repeats increased (p=0.63 and p=0.52, respectively) (**Figure 3C,D**). Age at enrollment did not moderate the variability in mean SBMAFRS scores (p=0.51) (**Figure 3A**).

**Figure 2.**
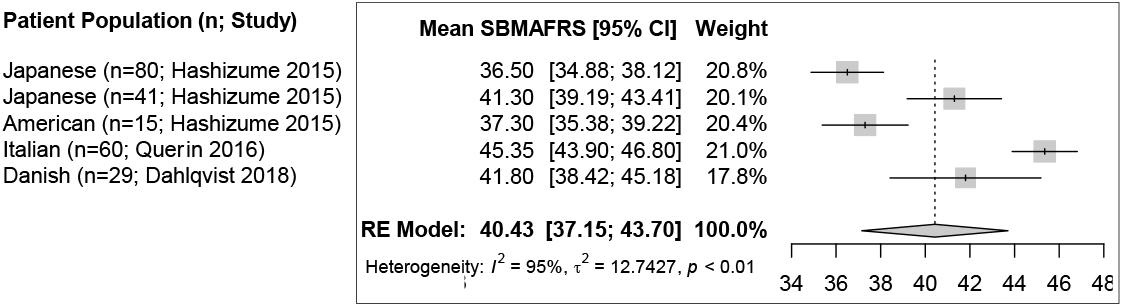
Forest plot of pooled mean SBMAFRS global score of five populations from three published studies.

**Figure 3.**
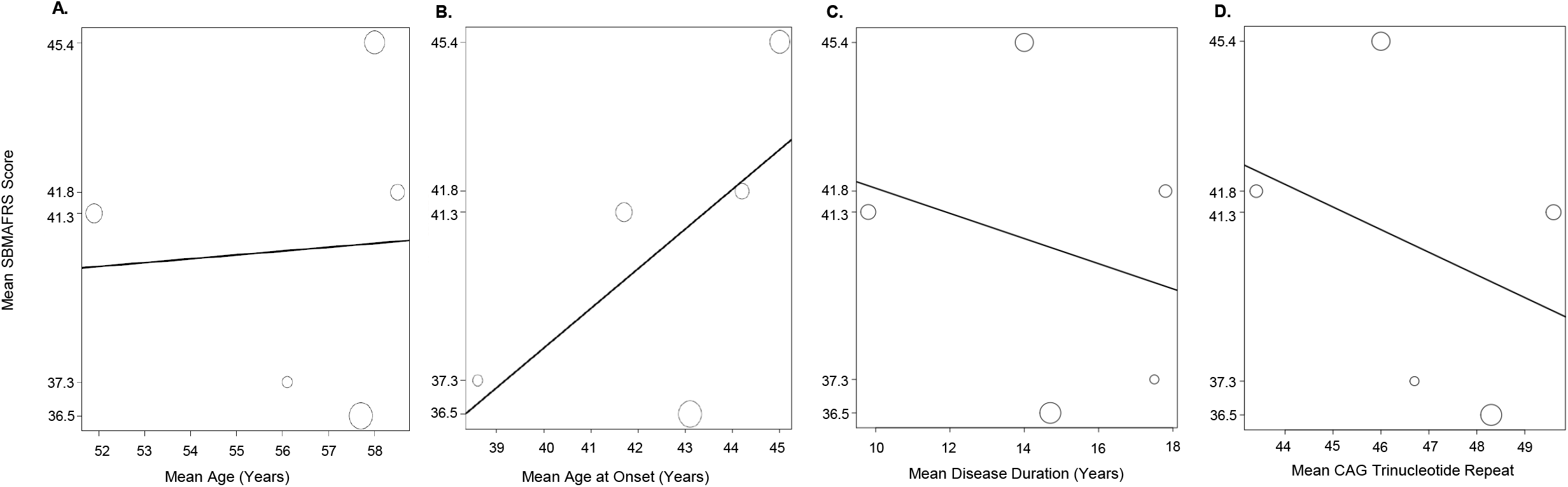
Meta-regression models of mean SBMAFRS scores across studies moderated to mean age at enrollment (A), mean age at onset (B), mean disease duration (C), and mean CAG trinucleotide repeats (D).

### 6. Purpose-specific Measurement Properties

Optimal intra- and inter-rater reliability, along with responsiveness, are paramount properties for the SBMAFRS to serve as an evaluative endpoint in multicentre clinical trials.

#### 5.1 Reliability studies

Intra- and inter-rater reliability measured by intraclass correlation coefficients (ICCs) were determined for each subscale and global scores of the SBMAFRS (**Table 1**). Intra-rater ICC was excellent for the global score (>0.9) as determined in Japanese patients (n=26). Inter-rater ICCs for global scores were also good (≥0.8) in Japanese (n=80) and American (n=15) patients. For most subscales and the global score, ICCs were above 0.7 except for the breathing subscale that had low inter-rater ICC (0.3) in the Japanese sample and high ICC (0.9) in the American population. This limitation may be related to inadequate response anchoring, as patients with early respiratory impairment could have discrepant scores depending on fluctuation or fatigability of the symptoms, or interpretation bias.

SBMAFRS test-retest reliability was also determined in the Italian population (n=60) using Spearman’s rho correlation and the percentage of perfect agreement between the test and retest [13]. Rho and percentage of perfect agreement were above 0.7 for all domains, except for breathing as seen in the Japanese study. These methods have weaknesses when compared to more standard methods such as ICC and kappa coefficients, as they do not account for variance matrix and the possibility of agreement occurring by chance alone, respectively.

Overall, the SBMAFRS was reliable, despite the poor inter-rater performance of the breathing item. Given respiratory impairment is an uncommon and late feature in SBMA, the measurement error of the breathing item may impact the SBMAFRS measures in patients with advanced disease.

#### 5.2 Longitudinal Construct Validity or Responsiveness

Based on the conceptual framework of tracking SBMA-specific motor functional loss over time, responsiveness to change is the key attribute to justify utilization of the SBMAFRS in clinical trials. The 3-axis taxonomy for responsiveness proposed by Beaton et al. 2001 [24] was used to establish, *a priori*, what to expect for the SBMAFRS responsiveness. Since prior natural history studies have revealed that SBMA motor function decline is reasonably linear and slow over time [25], observed changes in motor function (the “What Axis”) over a short interval of 12 months (the “Which Axis”) is likely of small magnitude. This assumption is supported by the mean SBMAFRS scores that was not significantly moderated by disease duration as noted by the meta-regression modelling (**Figure 3C**).

The SBMAFRS responsiveness over 48 weeks, as compared to the Limb Norris Score, Norris Bulbar Score, and ALSFRS-R, was evaluated in 41 Japanese patients aged 51.9±11.2 years, with disease duration of 9.8±5.7 years. The design of this prospective cohort study fit well with our *a priori* framework, but it was likely underpowered based on our sample estimation. Group-level data was presented as mean longitudinal change ± standard error (SE) and standardized response mean (SRM); where SE is a measure of the signal only, while SRM is a ratio of signal (mean change) over noise (standard deviation of the change). SRM allows for the comparison of sensitivity to change among different instruments, in which values of 0.2, 0.5, and over 0.8 suggest small, moderate and large magnitude of change respectively [26]. Raw, individual-level, SBMAFRS global and single item scores were plotted for the baseline and 48- week visits in this study. Longitudinal change at both individual- and aggregate-level are meaningful to clinical decision making, and determining sample size or outcome measures, respectively.

The responsiveness of the SBMAFRS was unsatisfactory for our measurement concept. The mean SBMAFRS global score change over 48 weeks was −1.27±0.44 point with an SRM of −0.48, while the mean ALSFRS-R change was −0.73±0.31 (SRM −0.39). Although the sensitivity of the SBMAFRS was superior to ALS-related functional scales, the detected magnitude of change in this underpowered study was too small and may be related to measurement error only. Additionally, patient-level plots revealed that a number of patients had stable SBMAFRS scores over time, which could be attributed to true, transient disease stability or lack of SBMAFRS sensitivity to mild changes.

## 4. DISCUSSION

SBMA is an incurable, slowly progressive, X-linked neurodegenerative disease. Many randomized, controlled clinical trials in SBMA have failed to demonstrate a therapeutic effect compared to placebo when ALS-related scales or unidimensional clinimetric measures are used as outcomes [4, 8, 19]. Besides the possibility of inefficacious therapy, negative trial results could be related to the lack of an instrument’s sensitivity to change (type 2 error). Thus, a SBMA-specific, valid, and responsive functional scale is needed for the success of upcoming clinical trials for promising therapeutics.

The SBMA Functional Rating Scale (SBMAFRS), published in 2015, has encouraging sensibility and feasibility to target our concept of tracking functional motor loss over time. For this scale to be used as an outcome measure in multicentre trials and registry-based or cohort studies however, its preliminary responsiveness (and possibly, its reliability) is demonstrated to be inadequate for measuring changes of small magnitude and could potentially jeopardize the detection of a possible small therapeutic effect in short-term trials.

Efforts should be made to improve the measurement properties of the SBMAFRS. One approach would be to reassess these parameters in a larger sample and/or lengthen a trial’s follow-up period to increase the statistical power. However, the generalizability of this approach and its implementation within feasible SBMA trials would be limited given the logistical challenges of running large, lengthy studies in rare disease such as SBMA. Another possibility would be to improve the scale’s sensitivity in detecting mild functional change within 6 to 12 months by modification of the response categories. This process would imply a return to the development stage, whereby SBMA experts, stakeholders and patients would revise the proposed subscales and judge meaningful edits or addition of new items supported by extensive data acquired during the original SBMAFRS validation phase. A major shortcoming of this conservative approach is halting therapeutic development by the additional expenses and lengthy instrument development process.

More recently, another SBMA-specific functional scale was published by Lu et al. – the Kennedy’s Disease 1234 scale (KD1234) [14]. This scale was developed by a single centre and validated in 81 Chinese patients. Like the SBMAFRS, the KD1234 has ten elements rated 0 to 3 spanning four domains (bulbar, upper limbs, lower limbs, and breathing). Although it has not yet been applied elsewhere, both the Cronbach’s alpha internal consistency and inter-rater reliability scored above 0.7, suggesting it may be feasibily implemented across multiple centres. Remarkably, the KD1234 breathing item with only four categories had relatively better reliability compared to the SBMAFRS breathing item [14]. Some KD1234 items probe intense and laborious motor activities such as push-ups, running, and squatting to standing positions, that may be more responsive to early functional loss. The responsiveness of the KD1234 over 32 months was suboptimal, with an SRM of −0.612 (n=52), also indicating a need for modification.

Given the shortcomings of both the SBMAFRS and KD1234, a more pragmatic approach to improve both scales could be implemented. Simultaneous administration of both the SMBAFRS and KD1234 in a large, multicentre, prospective cohort of SBMA patients could be conducted to assess their repeated measures and comparative performance. Both scales should be anchored to global indicators of change at baseline, 6 and 12 months with proper, videotaped rater training from standardized patients to strengthen instrument reproducibility. This strategy would allow for head-to-head comparison of the reliability and responsiveness while generating additional data to support the possible development of a combined scale if it is deemed a better alternative.

After carrying out the proposed multicentre cohort study, an expanded, combined scale could be statistically and judgmentally explored further. Grouping the KD1234 simplified breathing item, the additional laborious motor function items, and the most responsive SBMAFRS items may potentially help overcome the narrow responsiveness over shorter intervals, along with enhanced interpretability of disease progression versus stability as indicated by change in scale scores. Finally, parallel studies using the combined scale with SBMA-related wet, molecular, and neurophysiological biomarkers during the validation process may offer a harmonized measure to be used in short-term clinical trials to ultimately propel treatment advances in SMBA.

## 5. CONCLUSION

In summary, the SBMAFRS has satisfactory internal consistency, inter-rater reliability (except for the breathing item), and construct validity. Longitudinally, the SBMAFRS had superior responsiveness performance over existing ALS motor function scales but fell short in detecting considerable magnitude of progression-related change in an underpowered, preliminary cohort [11, 13]. Efforts to increase the SBMA Functional Rating Scale’s sensitivity to change are warranted and could be achieved by comparing and possibly combining the KD1234 with the SBMAFRS in a larger cohort study.

## Data Availability

Data referred to in this manuscript with be made available upon request to the corresponding author.

## DECLARATIONS OF INTEREST

The authors have no conflicts of interest to declare.

## APPENDIX.

**Supplementary Data**

## Notes

### Competing Interest Statement

The authors have declared no competing interest.

### Funding Statement

This article was supported by the intramural funds of the Sunnybrook ALS Research Program.

### Author Declarations

This article does not contain any studies with human participants performed by any of the authors.

