## Supplementary Tables and Figures for "Functional Rating Scales in Spinal and Bulbar Muscular Atrophy: A Systematic Review, Meta-Analysis and Critical Appraisal of their Measurement Properties"

**Supplementary Table 1.** Search Strategy

| **Database Name** | **PubMed** | **OVID Medline and Embase** |
| --- | --- | --- |
| **Search terms** | ((((((((((Kennedy's disease[MeSH] OR kennedy's disease[tiab] OR spinal and bulbar muscular atrophy[tiab] OR spinal bulbar muscular atrophy[tiab]))) AND (((functional[Title/Abstract] OR function[Title/Abstract] OR weakness[Title/Abstract] OR loss of function[Title/Abstract] OR symptom[Title/Abstract] OR disease progression[Title/Abstract] OR functional loss[Title/Abstract])))) AND ((scale[Title/Abstract] OR index[Title/Abstract] OR questionnaire[Title/Abstract] OR instrument[Title/Abstract]))))) AND ((instrumentation[sh] OR methods[sh] OR "Validation Studies"[pt] OR "Comparative Study"[pt] OR "psychometrics"[MeSH] OR psychometr*[tiab] OR clinimetr*[tw] OR clinometr*[tw] OR "outcome assessment (health care)"[MeSH] OR "outcome assessment"[tiab] OR "outcome measure*"[tw] OR "observer variation"[MeSH] OR "observer variation"[tiab] OR "Health Status Indicators"[Mesh] OR "reproducibility of results"[MeSH] OR reproducib*[tiab] OR "discriminant analysis"[MeSH] OR reliab*[tiab] OR unreliab*[tiab] OR valid*[tiab] OR "coefficient of variation"[tiab] OR coefficient[tiab] OR homogeneity[tiab] OR homogeneous[tiab] OR "internal consistency"[tiab] OR (cronbach*[tiab] AND (alpha[tiab] OR alphas[tiab])) OR (item[tiab] AND (correlation*[tiab] OR selection*[tiab] OR reduction*[tiab])) OR agreement[tw] OR precision[tw] OR imprecision[tw] OR "precise values"[tw] OR test-retest[tiab] OR (test[tiab] AND retest[tiab]) OR (reliab*[tiab] AND (test[tiab] OR retest[tiab])) OR stability[tiab] OR interrater[tiab] OR inter-rater[tiab] OR intrarater[tiab] OR intra-rater[tiab] OR intertester[tiab] OR inter-tester[tiab] OR intratester[tiab] OR intra-tester[tiab] OR interobserver[tiab] OR inter-observer[tiab] OR intraobserver[tiab] OR intra-observer[tiab] OR intertechnician[tiab] OR inter- technician[tiab] OR intratechnician[tiab] OR intra-technician[tiab] OR interexaminer[tiab] OR inter- examiner[tiab] OR intraexaminer[tiab] OR intra-examiner[tiab] OR interassay[tiab] OR inter-assay[tiab] OR intraassay[tiab] OR intra-assay[tiab] OR interindividual[tiab] OR inter-individual[tiab] OR intraindividual[tiab] OR intra-individual[tiab] OR interparticipant[tiab] OR inter-participant[tiab] OR intraparticipant[tiab] OR intra-participant[tiab] OR kappa[tiab] OR kappa's[tiab] OR kappas[tiab] OR repeatab*[tw] OR ((replicab*[tw] OR repeated[tw]) AND (measure[tw] OR measures[tw] OR findings[tw] OR result[tw] OR results[tw] OR test[tw] OR tests[tw])) OR generaliza*[tiab] OR generalisa*[tiab] OR concordance[tiab] OR (intraclass[tiab] AND correlation*[tiab]) OR discriminative[tiab] OR "known group"[tiab] OR "factor analysis"[tiab] OR "factor analyses"[tiab] OR "factor structure"[tiab] OR "factor structures"[tiab] OR dimension*[tiab] OR subscale*[tiab] OR (multitrait[tiab] AND scaling[tiab] AND (analysis[tiab] OR analyses[tiab])) OR "item discriminant"[tiab] OR "interscale correlation*"[tiab] OR error[tiab] OR errors[tiab] OR "individual variability"[tiab] OR "interval variability"[tiab] OR "rate variability"[tiab] OR (variability[tiab] AND (analysis[tiab] OR values[tiab])) OR (uncertainty[tiab] AND (measurement[tiab] OR measuring[tiab])) OR "standard error of measurement"[tiab] OR sensitiv*[tiab] OR responsive*[tiab] OR (limit[tiab] AND detection[tiab]) OR "minimal detectable concentration"[tiab] OR interpretab*[tiab] OR ((minimal[tiab] OR minimally[tiab] OR clinical[tiab] OR clinically[tiab]) AND (important[tiab] OR significant[tiab] OR detectable[tiab]) AND (change[tiab] OR difference[tiab])) OR (small*[tiab] AND (real[tiab] OR detectable[tiab]) AND (change[tiab] OR difference[tiab])) OR "meaningful change"[tiab] OR "ceiling effect"[tiab] OR "floor effect"[tiab] OR "Item response model"[tiab] OR IRT[tiab] OR Rasch[tiab] OR "Differential item functioning"[tiab] OR DIF[tiab] OR "computer adaptive testing"[tiab] OR "item bank"[tiab] OR "cross- cultural equivalence"[tiab]))) NOT (("addresses"[Publication Type] OR "biography"[Publication Type] OR "case reports"[Publication Type] OR "comment"[Publication Type] OR "directory"[Publication Type] OR "editorial"[Publication Type] OR "festschrift"[Publication Type] OR "interview"[Publication Type] OR "lectures"[Publication Type] OR "legal cases"[Publication Type] OR "legislation"[Publication Type] OR "letter"[Publication Type] OR "news"[Publication Type] OR "newspaper article"[Publication Type] OR "patient education handout"[Publication Type] OR "popular works"[Publication Type] OR "congresses"[Publication Type] OR "consensus development conference"[Publication Type] OR "consensus development conference, nih"[Publication Type] OR "practice guideline"[Publication Type]) NOT ("animals"[MeSH Terms] NOT "humans"[MeSH Terms])))) | 1. "spinal and bulbar muscular atrophy".mp. or exp Kennedy disease/  2. (instrumentation or methods).sh.  3. (Validation Studies or Comparative Study).pt.  4. exp Psychometrics/  5. psychometr*.ti,ab.  6. (clinimetr* or clinometr*).tw.  7. exp outcome assessment/  8. outcome assessment.ti,ab.  9. outcome measure*.tw.  10. exp Observer Variation/  11. observer variation.ti,ab.  12. exp Health Status Indicators/  13. exp reproducibility/  14. reproducib*.ti,ab.  15. exp Discriminant Analysis/  16. reliab*.ti,ab.  17. unreliab*.ti,ab.  18. valid*.ti,ab.  19. coefficient.ti,ab.  20. homogeneity.ti,ab.  21. homogeneous.ti,ab.  22. internal consistency.ti,ab.  23. (cronbach* and (alpha or alphas)).ti,ab.  24. (item and (correlation* or selection* or reduction*)).ti,ab.  25. agreement.ti,ab.  26. precision.ti,ab.  27. imprecision.ti,ab.  28. precise values.ti,ab.  29. test-retest.ti,ab.  30. (test and retest).ti,ab.  31. (reliab* and (test or retest)).ti,ab.  32. stability.ti,ab.  33. interrater.ti,ab.  34. inter-rater.ti,ab.  35. intrarater.ti,ab.  36. intra-rater.ti,ab.  37. intertester.ti,ab.  38. inter-tester.ti,ab.  39. intratester.ti,ab.  40. intra-tester.ti,ab.  41. interobserver.ti,ab.  42. inter-observer.ti,ab.  43. intraobserver.ti,ab.  44. intertechnician.ti,ab.  45. inter-technician.ti,ab.  46. intratechnician.ti,ab.  47. intra-technician.ti,ab.  48. interexaminer.ti,ab.  49. inter-examiner.ti,ab.  50. intraexaminer.ti,ab.  51. intra-examiner.ti,ab.  52. interassay.ti,ab.  53. intra-observer.ti,ab.  54. inter-assay.ti,ab.  55. intraassay.ti,ab.  56. intra-assay.ti,ab.  57. interindividual.ti,ab.  58. inter-individual.ti,ab.  59. intraindividual.ti,ab.  60. intra-individual.ti,ab.  61. interparticipant.ti,ab.  62. inter-participant.ti,ab.  63. intraparticipant.ti,ab.  64. intra-participant.ti,ab.  65. kappa.ti,ab.  66. kappas.ti,ab.  67. repeatab.ti,ab.  68. ((replicab* or repeated) and (measure or measures or findings or result or results or test or tests)).ti,ab.  69. (generaliza* or generalisa* or concordance).ti,ab.  70. (intraclass and correlation*).ti,ab.  71. discriminative.ti,ab.  72. known group.ti,ab.  73. factor analysis.ti,ab.  74. factor analyses.ti,ab.  75. dimension.ti,ab.  76. subscale.ti,ab.  77. (multitrait and scaling and (analysis or analyses)).ti,ab.  78. item discriminant.ti,ab.  79. interscale correlation*.ti,ab.  80. error.ti,ab.  81. errors.ti,ab.  82. individual variability.ti,ab.  83. (variability and (analysis or values)).ti,ab.  84. (uncertainty and (measurement or measuring)).ti,ab.  85. standard error of measurement.ti,ab.  86. sensitiv*.ti,ab.  87. responsive*.ti,ab.  88. ((minimal or minimally or clinical or clinically) and (important or significant or detectable) and (change or difference)).ti,ab.  89. (small* and (real or detectable) and (change or difference)).ti,ab.  90. meaningful change.ti,ab.  91. ceiling effect.ti,ab.  92. floor effect.ti,ab.  93. Item response model.ti,ab.  94. IRT.ti,ab.  95. Rasch.ti,ab.  96. Differential item functioning.ti,ab.  97. DIF.ti,ab.  98. computer adaptive testing.ti,ab.  99. item bank.ti,ab.  100. cross-cultural equivalence.ti,ab.  101. 2 or 3 or 4 or 5 or 6 or 7 or 8 or 9 or 10 or 11 or 12 or 13 or 14 or 15 or 16 or 17 or 18 or 19 or 20 or 21 or 22 or 23 or 24 or 25 or 26 or 27 or 28 or 29 or 30 or 31 or 32 or 33 or 34 or 35 or 36 or 37 or 38 or 39 or 40 or 41 or 42 or 43 or 44 or 45 or 46 or 47 or 48 or 49 or 50 or 51 or 52 or 53 or 54 or 55 or 56 or 57 or 58 or 59 or 60 or 61 or 62 or 63 or 64 or 65 or 66 or 67 or 68 or 69 or 70 or 71 or 72 or 73 or 74 or 75 or 76 or 77 or 78 or 79 or 80 or 81 or 82 or 83 or 84 or 85 or 86 or 87 or 88 or 89 or 90 or 91 or 92 or 93 or 94 or 95 or 96 or 97 or 98 or 99 or 100  102. 1 and 101 |

**Supplementary Figure 1.** PRISMA flow chart of articles selected for meta-analysis.

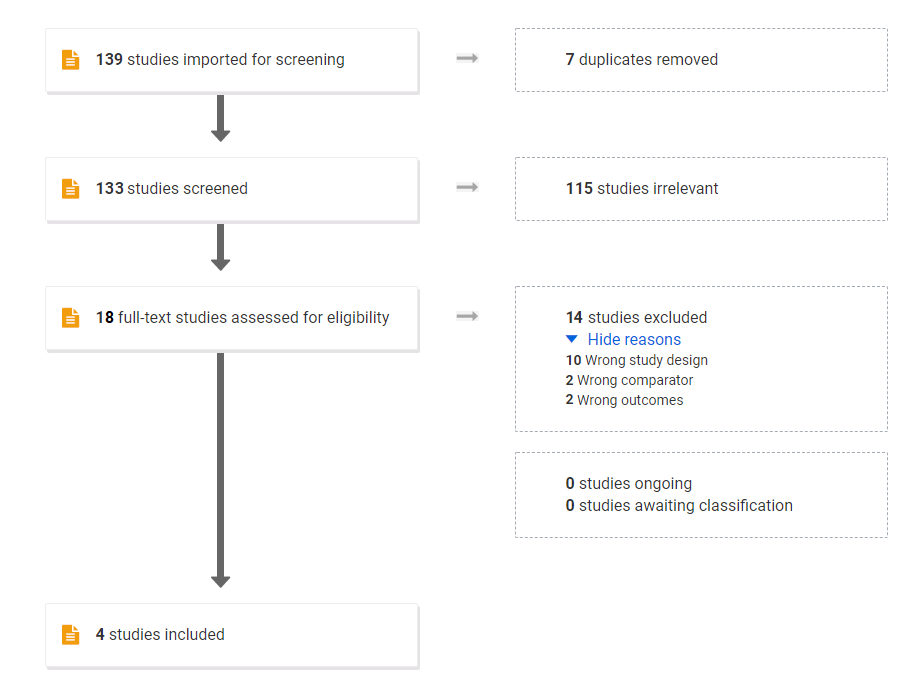
